# The cross-cultural validation of the Beach Center Family Quality of Life Scale among persons affected by leprosy and podoconiosis in Northwest, Ethiopia

**DOI:** 10.1101/2023.03.16.23287345

**Authors:** Moges Wubie Aycheh, Anna Tiny van ’t Noordende, Nurilign Abebe Moges, Alice Schippers

## Abstract

**Background:** The Beach Center Family Quality of Life Scale was developed and validated in different languages in different countries. However, this scale is not yet validated in the Ethiopian Amharic language context. Therefore, this study aimed to investigate the cross-cultural validity of the Beach Center Family Quality of Life Scale, among Ethiopian families of persons affected by leprosy and podoconiosis.

**Methodology:** We explored the semantic equivalence, internal consistency, reproducibility, floor and ceiling effects, and interpretability of the Beach Centre Family Quality of Life Scale in Amharic. After translating, and back-translating the instrument, a cross-sectional study was conducted. A total of 302 adult persons affected by leprosy and podoconiosis, who have a view of their family life were asked about their level of satisfaction using the Beach Center Family Quality of Life Scale. In addition, 50 participants were interviewed again two weeks after the first assessment to check the reproducibility of the scale. Participants were recruited in the East Gojjam zone in Northwest Ethiopia.

**Results:** The findings of this study showed that the Beach Center Family Quality of Life Scale had high internal consistency (Cronbach’s alpha of 0.913) and reproducibility (intra-class correlation coefficient of 0.857). The standard error of measurement was 3.01, which is 2.4% of the total score range. The smallest detectable change was 8.34. Confirmatory factor analysis showed adequate factor loadings and model fit indices like the original scale. The composite reliability and average variance extracted from the scale were acceptable. No floor and ceiling effects were found.

**Conclusions:** Our findings indicate that the Amharic version of the Beach Center Family Quality of Life scale has adequate cultural validity to assess the family quality of life in Ethiopian families of persons affected by leprosy and podoconiosis.

**Author Summary:** The presence of persons with disabilities in the family can affect a family’s quality of life. Neglected tropical diseases such as leprosy and podoconiosis can lead to disabilities and have been found to influence Family Quality of Life. To adequately assess the family quality of life of persons affected by leprosy and podoconiosis, we have selected the Beach Center Family Quality of Life scale. However, this scale has not been validated in the Ethiopian context previously. In this study, the authors aimed to investigate the cross-cultural validity of the Beach Center Family Quality of Life scale in families with one or more persons affected by leprosy and podoconiosis in Ethiopia.

A total of 302 participants were included in this study. The results show the scale to be adequately reliable and valid in the target country’s culture and language. Based on the findings, the authors recommend using this scale among families of persons affected by leprosy and podoconiosis.

## Introduction

Family Quality of Life (FQoL) is an extension of individual quality of life (QoL) [1-3]. Recently, the concept of FQoL has developed out of the broader quality of life work and has been applied to families in which a member has an intellectual disability [4], and in families with members with other conditions such as Alzheimer’s disease, serious medical conditions, and families that live in disadvantaged communities [5]. FQoL is a multidimensional construct [3, 6, 7] and is defined as “conditions where the family’s needs are met, and family members enjoy their life together as a family and have the chance to do things which are important to them” [8]. Using empirical data from their own literature review, Zuna et al proposed four main concepts that influence variations in FQoL: (i) systemic concepts (systems, policies, and programs); (ii) performance concepts (services, supports, and practices); (iii) individual-member concepts (demographics, characteristics, and beliefs); and (iv) family-unit concepts (characteristics, and dynamics) [3]. Zuna et al four combined concepts changed to practices of interventions aimed at improving individual quality of life. Such an approach is more effective when we also take the family, culture, and environmental context into account [4, 9-12]. FQoL is therefore one of the main outcome measures of services and the provision of family support for people with intellectual disabilities [7, 9, 13]. Guidance from family members is helpful for individuals with disabilities [14-17]. In this sense, in the past two decades, the FQoL concept received greater attention for its development and utilization [9, 18], especially in the field of disability studies [19, 20].

FQoL has been assessed using qualitative [6, 10] and quantitative [10-12] approaches. Especially for the latter approach, scholars have developed different scales to measure FQoL, such as the Beach Center Family Quality of Life Scale (BC-FQoL) [11], the International Family Quality of Life scale [12], and a Latin America FQoL Scale [10]. Among these, we selected the BC-FQoL scale [11] to measure the FQoL for families of persons with leprosy and podoconiosis-related disabilities. Because this scale is one of the most used instruments to assess FQoL in families with special needs. It provides concise and quick information about families⍰. overall well-being and can be used to address the needs of families giving help to persons with disabilities within their homes [16]. In addition, the BC-FQoL is a psychometrically comprehensive measure that can be used in research and clinical practice to evaluate programs and services for families of children with disabilities [21].

The BC-FQoL scale was originally developed in United States of America [11]. However, a scale developed in one country may not work in the same manner in another country [22]. Based on this concept, a culturally adaptable, valid, and reliable scale plays a pivotal role in the measurement of FQoL of persons with disabilities and their family members. Previously, the BC-FQoL Scale has been validated in Spanish [7, 9], Chinese [23, 24], Turkish [25], and (Brazilian) Portuguese [26]. The BC-FQoL Scale has not been cross-culturally validated in the Ethiopian context. Therefore, there is a need to culturally validate the scale in Ethiopia before using it to measure the FQoL persons affected by leprosy and podoconiosis.

Therefore, the aim of this study was to investigate the cross-cultural validity of the BC-FQoL scale among persons affected by leprosy and podoconiosis in Northwest Ethiopia. Furthermore, we also aimed to conduct a cross-cultural adaptation of the BC-FQoL Scale to the Amharic language.

## Methodology

### Ethical considerations

This study was approved by the Debre Markos University, Health Sciences College, Institutional Research Ethics Review Committee (IRERC) with reference number HSR/R/C/Se/Co/11/13. In addition, permission letter obtained from the Amhara Public Health Institute. Both the nature and objective of the study and the confidentiality of the data was clarified to each study participant before the data collection. Participation in the study was voluntarily. Because of the low literacy level among the study participants, participants gave their verbal informed consent.

### Study design

The study was a cross-sectional scale validation study.

### Study site

This study was conducted in Northwest Ethiopia in the East Gojjam Zone. East Gojjam, also called Misraq Gojjam, is a zone in the Amhara Region of Ethiopia. The zone’s capital city is Debre Markos. Debre Markos is located 300 km away from Addis Ababa, Ethiopia. It is bordered in the South by the Oromia Region, the West-by-West Gojjam, in the North by South Gondar, and in the East by South Wollo. The east Gojjam zone has a total population of 2,719,118 people, which comprise of 632,353 households. The zone also has 21 woredas (districts), 480 kebeles (the smallest administrative unit), 423 health posts, 102 health centers, 10 primary hospitals, and one referral hospital [27]. The main language is Amharic. Both leprosy and podoconiosis are prevalent in the area [28].

### Study population, sample size and sampling technique

For the validation of the BC-FQoL, different population groups were selected for the collection of data. Out of the research team members, six experts participated in the translation, back translation, and evaluation process. Six persons with podoconiosis also participated for checking the completeness, understandability of the scale as part of the validation of the study. To ensure quantitative validation, we sought to include 300 persons affected by leprosy and podoconiosis with disabilities who are adults and have a view of their family life. Different scholars recommended that the sample size should be calculated in different ways. Based on Terwee et al 7:1 [29], Kline recommend a participant-to-indicator ratio of 10 up to 20:1 [30], and Viswesvaran 15:1 or 30:1 [31]. However, according to Comfrey and his colleagues, a sample size of 50 is considered very poor, 100 is considered poor, 200 is considered fair, 300 is considered good, 500 is considered very good, and 1000 or more as excellent [32]. Based on their recommendations, we opted to use a sample size of 300 on the basis that it would be sufficiently accurate for our needs. Participants were selected based on convenience sampling. In addition, 50 samples were randomly selected from the initially sampled population. These people were interviewed again two weeks after the original interviews were conducted as a means of checking the consistency of the scale used.

### Eligibility criteria

For the translation, evaluation and back translation of the BC-FQoL Scale researchers, language experts, psychologist, mental health experts and one of the original scale developer participated. For quantitative validation, participants who had to live in one of the five districts included in the study. The persons affected had to be diagnosed with leprosy or podoconiosis and had to have visible impairments due to their condition. Persons unwilling or unable to give verbal informed consent, persons younger than 16 years of age were excluded.

### Measures

The BC-FQoL scale was originally developed by researchers at the Beach Centre on Disability, a research and training center of the University of Kansas [11]. The scale is intended to quantify the insights and levels of satisfaction persons with disabilities experience within their Family Quality of Life [8]. This scale contains 25 items with a 5-point Likert scale (1-very dissatisfied, 2-dissatisfied, 3-neither satisfied nor dissatisfied, 4-satisfied, and 5-very satisfied). The scale consists of five subscales: 1) family interaction (6 items), 2) parenting (6 items), 3) emotional well-being (4 items), 4) physical/material well-being (5 items) and 5) disability-related support (4 items). The total scores for the satisfaction ratings range from 25 to 125 [11]. In addition to the scale sociodemographic information collected.

### Translation and adaptation process

We adapted the original English version of the BC-FQoL to Amharic using the procedure outlined by Borsa et al. [33]. The cross-cultural validation of the BC-FQoL measurement scale consisted of two main phases. The first phase entailed translation and adaptation, which itself has five stages: 1) instrument translation from the source language (English) into the target language (Amharic), 2) synthesis of the translated content, 3) a synthesis evaluation by experts, 4) tool evaluation by the target population (persons with leprosy and podoconiosis), and 5) back translation into English language. In addition, different equivalence of the scale and its subscales was assessed based on different definitions and criteria such as the conceptual equivalence pursued through a rigorous process, including forward and backward translation. Item equivalence considered by the degree to which the items composing the instrument are identical across cultures. Operational equivalence refers to the possibilities of using a similar questionnaire format, instructions, mode of administration and measurement methods [29, 34, 35]. The second phase entailed a quantitative validation (assessment of measurement equivalence).

For the initial step of the first phase, the English version of the BC-FQoL Scale was translated by two authors whose mother tongue is Amharic. First, the researchers performed the translation independently and then the two translated versions were compared and discussed to ensure semantic equivalence and agreement with the conceptual framework of the original scale. In cases of disagreement between the two authors, a third person was invited to solve the disagreement. This was facilitated using three additional experts (a psychologist, an Amharic language professor, and a psychiatrist) whose mother tongue is also Amharic. Moreover, the three experts were also invited to assess the semantic equivalence of the translation and to provide the authors with written feedback.

Back-translation to English was performed by two professors who were fluent in English and Amharic. The back translation was also done independently and without disclosing the original version of the scale to the translators. The back-translated version was compared to the original scale by three authors, and small changes were consequently made on the back translated version of the scale. Finally, the translated and back-translated scale was sent to the original scale developers who were asked to review the translations. Based on their feedback, small corrections were made in the Amharic version of the scale.

Furthermore, six persons affected by podoconiosis were asked to assess the legibility, clarity, and cultural suitability of the Amharic version of the scale. These persons reported that the items reflected their ideas, and the wording of the items was clear and could be easily understood. The scale demonstrated adequate face validity. It took about 25 minutes per participant to complete the interviews with each of the six persons affected by podoconiosis.

### Data collection

Seven health professionals participated in the data collection process. A two-day training on the objective of the study and the details of the scale idea was given to them by two authors of this article. The data collectors contacted persons with leprosy and podoconiosis in their homes, and around their locality. This was done after contacting them through their kebele and leprosy association leaders. The data collectors explained the objectives of the study and obtained verbal informed consent before the start of the interview. Confidentiality of the data was guaranteed throughout the study. Participants were interviewed face to face at their home or around their locality in a quiet room. The data were collected from August – October 2021.

### Data analysis

Epi data version 3.1 was used for data entry, and SPSS version 25.0 for data analysis. SPSS AMOS version 21.0 was used for confirmatory factor analysis (CFA).

Different statistical analyses were carried out to determine the reliability of the Beach Centre FQoL scale as translated to Amharic. Cronbach’s alpha was calculated for the total score of the scale and the subscales to determine the internal consistency of the instrument. In addition, the test–retest procedure was used to evaluate the reliability and reproducibility of measures. This was done with a subsample comprising 50 participants from the overall sample and using a time interval of two weeks. The test-retest reliability was assessed using the intra-class correlation coefficient (ICC) and its corresponding 95% confidence interval. The internal consistency and test-retest reliability were considered acceptable when the values of Cronbach⍰.s alpha and ICC exceeded 0.70 [36, 37]. The standard error of measurement (SEMagreement) was also calculated to determine the reproducibility of the scale, using the formula SEM = SD * (√1-ICC). The SEM was also converted into the smallest detectable change (SDC=1.96 X √2 X SEM), which reflects the smallest within-person change in score. With a P<0.05, this can be interpreted as a ‘‘rea⍰’ change, above measurement error, in one individual (SDCind). The SDC measurable in a group of people (SDCgroup) can be calculated by dividing the SDCind by √n. Values above the SDC describe a change in the individua’s score above the error of the measurement [38, 39]. Floor and ceiling effects were also calculated. Floor and ceiling effects are considered present if more than 15% of the respondents achieved the lowest or highest possible score on the scale [39-41].

Finally, Confirmatory Factor Analysis (CFA) with the maximum likelihood estimation was performed to examine the dimensionality and construct validity of the five-factor structure of the BC-FQoL. When the first-order latent variables were mutually related and can be accounted by a second-order latent variable. We examined whether the second-order five-factor structure of the BC-FQoL fits the Ethiopian family context of persons affected by leprosy and podoconiosis. Model fit was assessed using fit indices including the ratio of χ2 to the degrees of freedom (the χ2/df ratio), the goodness-of-fit index (GFI), the adjusted goodness-of-fit index (AGFI), the root means square error of approximation (RMSEA), the comparative fit index (CFI) and the incremental fit index (IFI). A value of 0.90 or more for the CFI and IFI [42, 43], a value of less than 5 for χ2/df, a value of 0.80 or more for the GFI and AGFI, and an RMSEA value between 0.05 and 0.08 are considered good model fit [44]. In addition, construct reliability (composite reliability (CR)) of 0.70 or more, and average variance extracted (AVE) of 0.50 or more were used to assess the convergent validity [45].

## Results

### Semantic equivalence

Minor changes were made to the first version of the back translated English version of the scale. For example, for item number 2, originally it said, “My family members help the children learn to be independent.” Interpreter 1, translated this as “My family members support children to be self-dependent and responsible.” Interpreter 2 translated this as “My family members help children to know about self-reliance/management.” We have selected interpreter 2’s translation by avoiding management at the end of the translated statement. Similarly for item number 13, “My family has outside help available to us to take care of special needs of all family members.” However, interpreter 1 translated this as “There is another body/person to support my family in times of difficulty.” Interpreter 2 translated it as “We have another person or person to take care of the special needs of my family members.” Based on this we have selected the later translation.

### Socio-demographic characteristics of the study participants

A total of 302 persons affected by leprosy (n=166, 55%) or podoconiosis (n=136, 45%) participated in the study. Over half of the study participants were male (n=178, 59%), and three quarters of the study participants (n=226, 75%) were below 64 years of age (18 – 64 years, which is independent age segment of the population in Ethiopian context). The mean age of the study participants was 54 (±14.2) years. About 79% (n= 239) of the study participants had a family size of below five people (which is the average family size of Ethiopia 4.8). Almost 80% (n=241) of the participants were not able to read and write and most of the participants (n=265, 88%), were farmers. An overview of the demographic information of the participants can be found in Table 1. Table 1 also includes the demographic characteristics of the 50 participants that were interviewed again after two weeks.

**Table 1.**
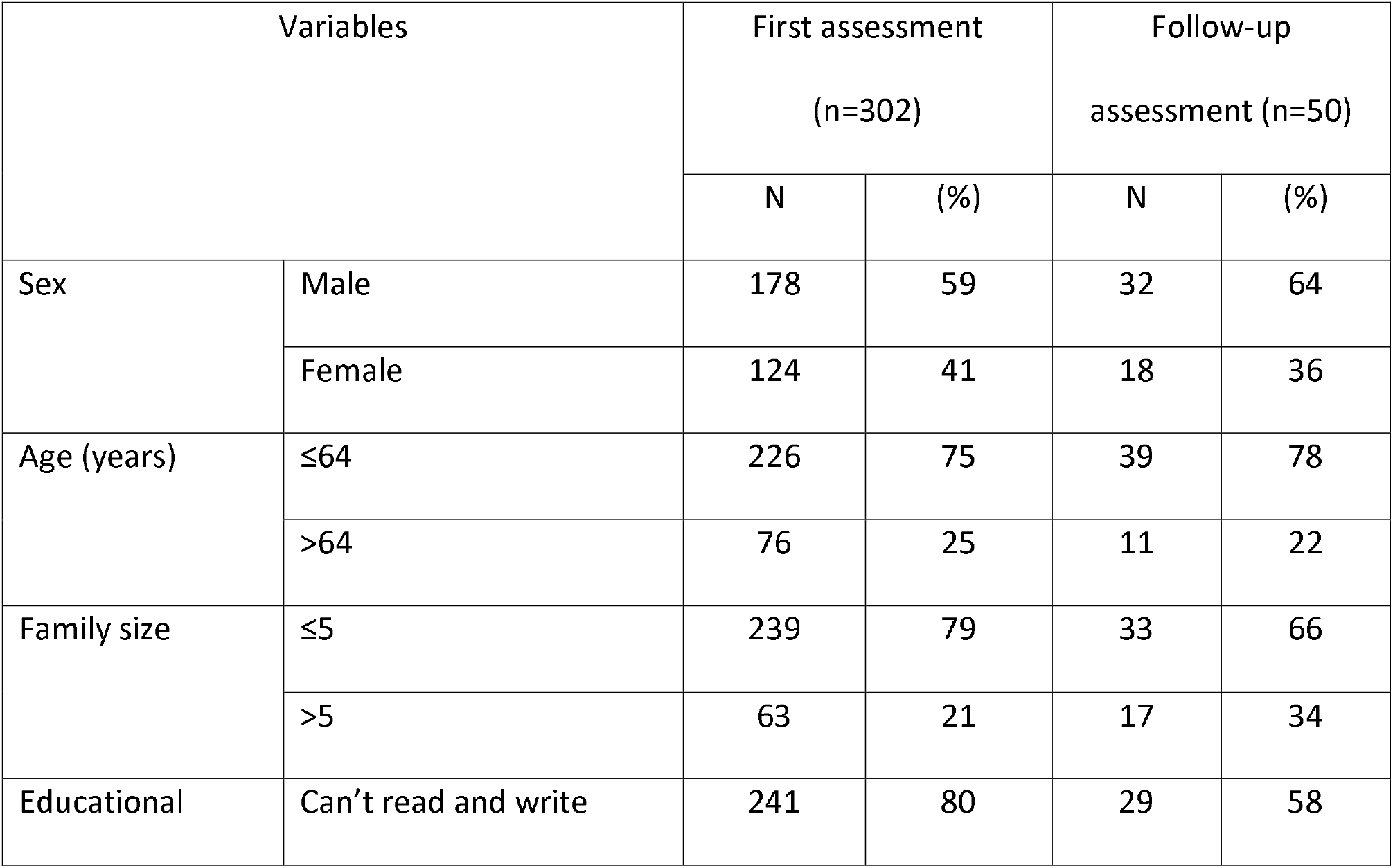
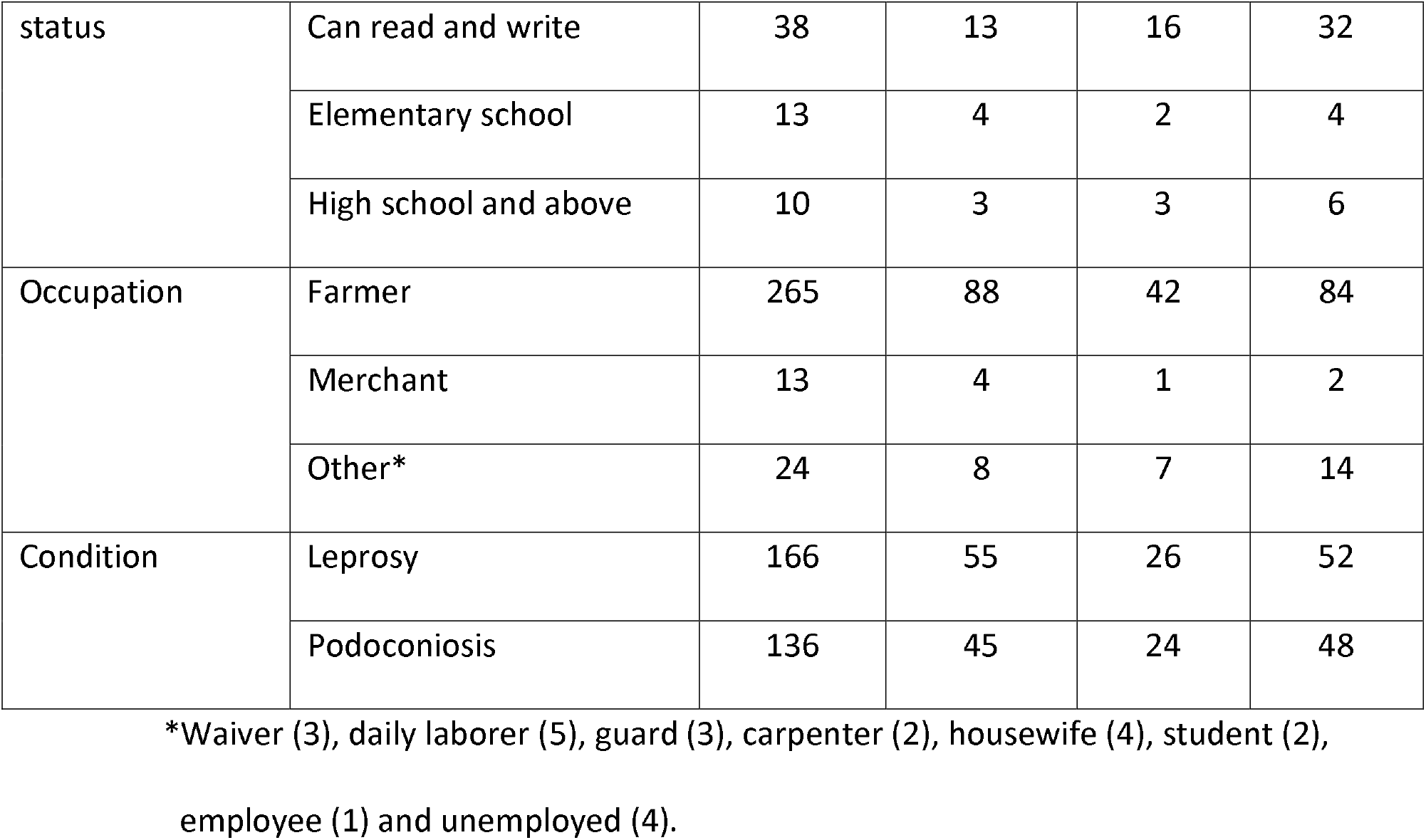
Sociodemographic characteristics of persons affected by leprosy and podoconiosis.

### Internal consistency

The internal consistency of the BC-FQoL Scale, the overall Cronbach’s alpha, was 0.913. For the five subscales, Cronbach’s alpha ranged from 0.683 to 0.850. Additionally, different items were deleted and calculated the Cronbach alpha. However, there was no visible difference in Cronbach alpha of both the subscales and the overall BC -FQoL Scale. Reliability statistics are summarized in Table 2.

**Table 2.** Reliability of the translated BC-FQoL scale.

| Subscales | No.<br>of<br>items | Cronbach's<br>alpha (n =<br>302) | Cronbach's<br>alpha (if<br>item is<br>deleted)<br>(n = 302) | ICCagreement<br>(95% CI) (n = 50) | SEM<br>(n =<br>50) | SEM<br>as %<br>of<br>total<br>score<br>range | SDC<sub>ind</sub><br>= SDC<br>(n = 50) | SDC<br>group<br>(n =<br>50) |
| --- | --- | --- | --- | --- | --- | --- | --- | --- |
| Family<br>interaction | 6 | 0.779 | 0.803 (N18) | 0.613(0.404, 0.760) * | 2.35 | 7.8% | 6.51 | 0.92 |
| Parenting | 6 | 0.692 | 0.697 (N17) | 0.578(0.362, 0.736) * | 2.56 | 8.5% | 7.10 | 1.00 |
| Emotional well-being | 4 | 0.683 | 0.708 (N9) | 0.508(0.268, 0.688) * | 1.57 | 7.9% | 4.35 | 0.62 |
| Physical/material<br>well-being | 5 | 0.728 | 0.718 (N21) | 0.618(0.411, 0.764) * | 1.95 | 7.8% | 5.41 | 0.76 |
| Disability-related<br>support | 4 | 0.850 | 0.842 (N24) | 0.847(0.744, 0.910) * | 0.92 | 4.6% | 2.55 | 0.36 |
| FQoL total score | 25 | 0.913 | 0.913 (FI) | 0.857(0.761,0.916) * | 3.01 | 2.4% | 8.34 | 1.18 |

\*P < 0.001, N represent item number of the BC-FQoL Scale

### Reproducibility: reliability and agreement

The BC-FQoL scale total score ICCagreement was 0.857 (95% CI: 0.761 – 0.916, p < 0.001). For the five subscales the ICCagreement ranged from 0.508 – 0.847. The subscale ICCagreement is summarized in Table 2. The standard error of measurement (SEM) was 3.01, this is 2.4% of the total score range. The smallest detectable change (SDC) was 8.34, and the SDC group 1.18. Details are summarized in Table 2.

### Validity

The five-factor CFA model yielded an acceptable model fit and the five first-order latent variables correlated well with each other, 0.73–0.91; all p < 0.001 (Fig. 1). Therefore, we conducted a second-order CFA model to examine the validity of the BC-FQoL (Fig. 2). The standardized factor loadings were all significant (p < 0.001), ranging from 0.65 to 0.99 ratings. Items all loaded well on the expected latent constructs. The results suggested that improving the model fit indices yielded an almost adequate fit (the χ2/df ratio 2.941; GFI .820; AGFI .791; IFI .818; CFI .817 and RMSEA 0.08). This result came after removing item number 5 from the parenting subscale, item number 9 from emotional well-being subscale and item number 25 from disability related support subscale. The CR values were all well above 0.70 for the satisfaction ratings. The AVE values which satisfied the criteria of 0.50 and above for all the subgroups of the BC-FQoL, except the parenting which was almost on the margin of 0.5 (0.492). Details can be found in Table 3.

**Table 3.** The subgroup of BC-FQoL composite reliability and average variance extracted.

|  | Composite reliability | Average Variance Extracted |
| --- | --- | --- |
| Family interaction | 0.896 | 0.592 |
| Disability related support | 0.884 | 0.658 |
| Physical/material well-being | 0.841 | 0.516 |
| Emotional well-being | 0.802 | 0.504 |
| Parenting | 0.852 | 0.494 |

**Figure 1.**
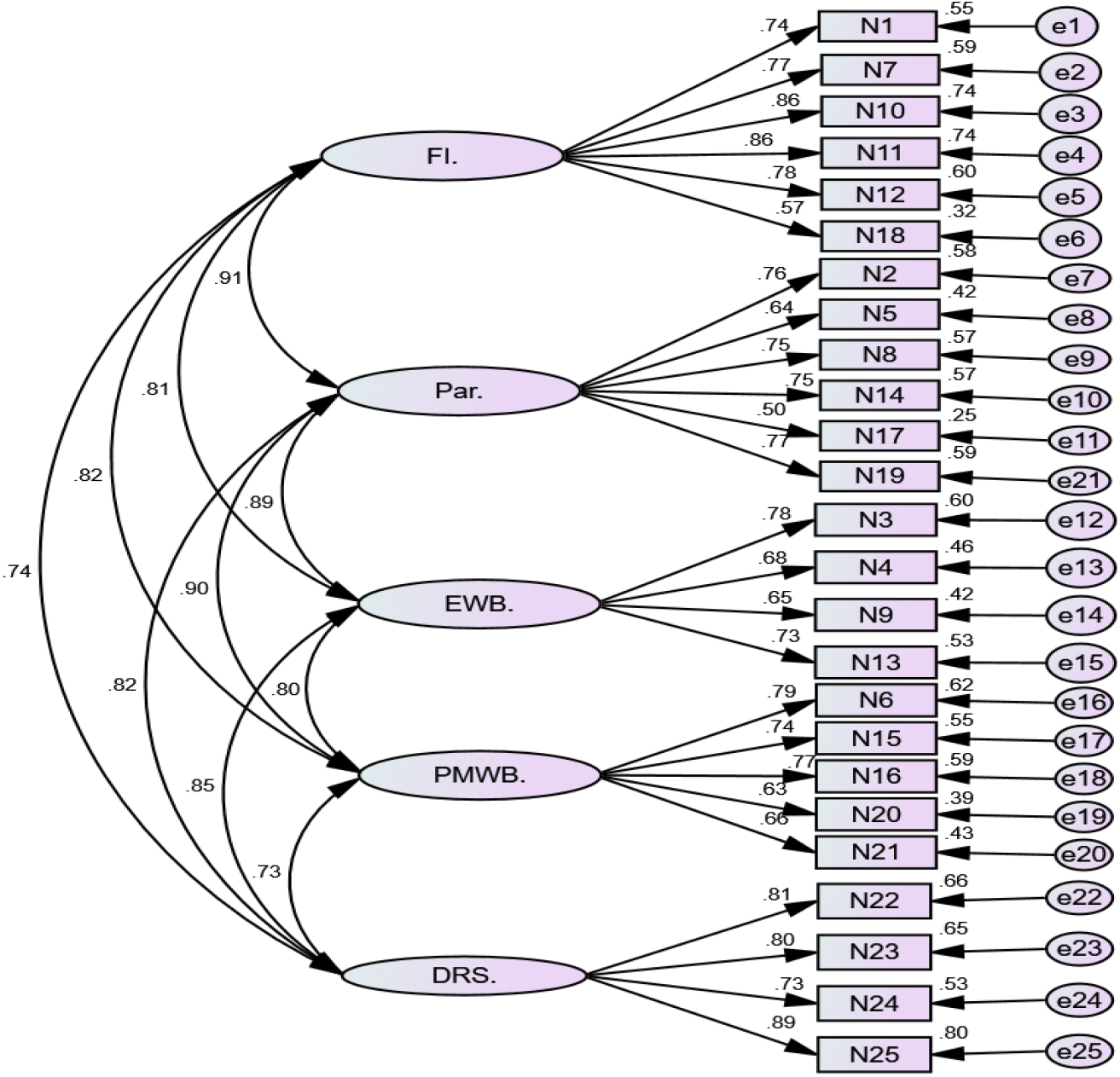
Measurement model of the Beach Centre Family Quality of Life scale and its result.

**Figure 2.**
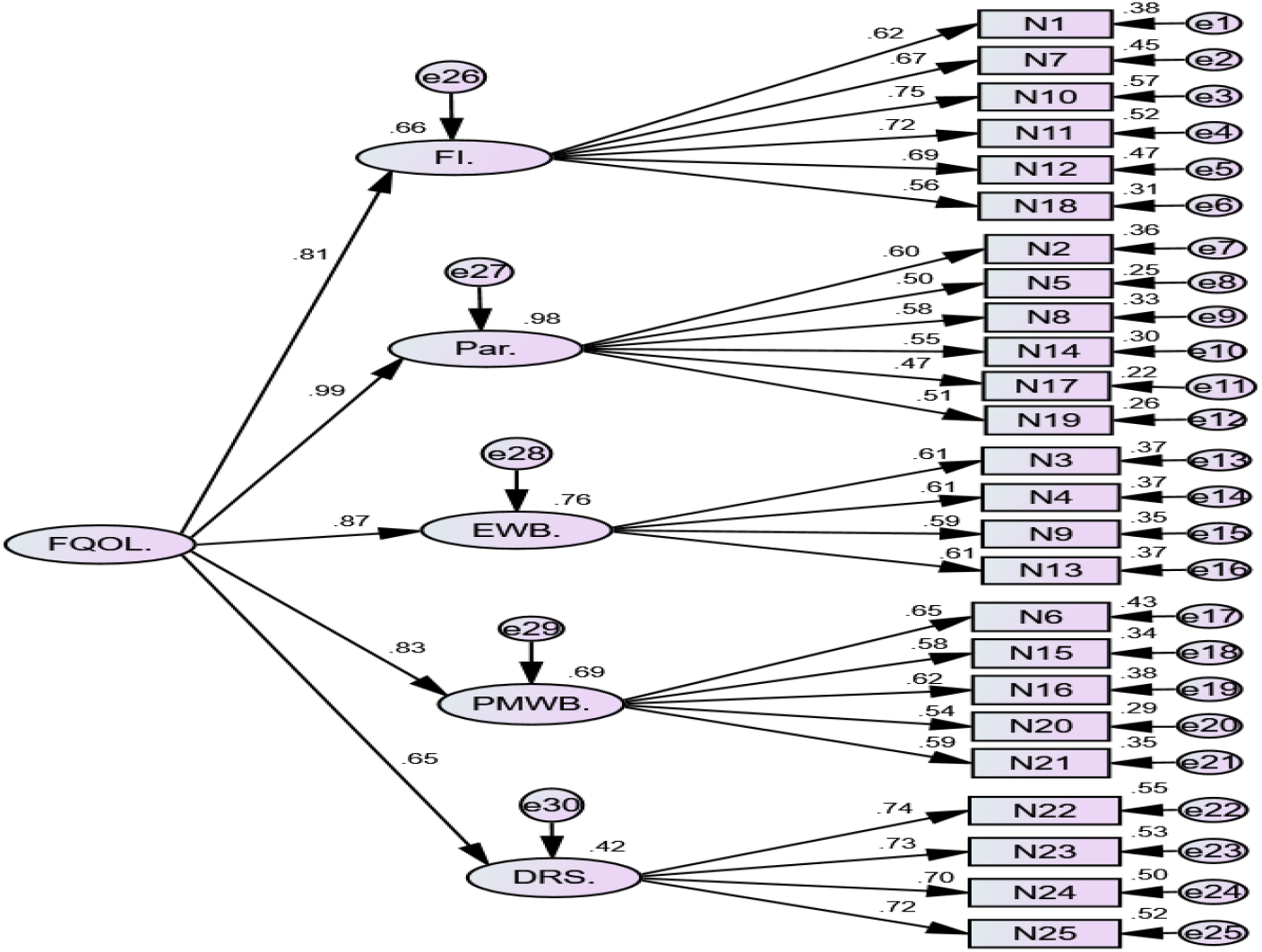
Standardized parameter estimates from confirmatory factor analysis of the Beach Centre Family Quality of Life scale: second-order five-factor model.

### Floor and ceiling effects

There are no floor and ceiling effects (Table 4).

**Table 4.** Floor and ceiling effect of BC-FQoL scale among persons affected by leprosy and podoconiosis.

| (Sub)scale | Range of (sub)scale | Number of people with the lowest score | Floor effect* | Number of people with the highest score | Ceiling effect* |
| --- | --- | --- | --- | --- | --- |
| Family interaction | 6-30 | 1 | No | 15 | No |
| Parenting | 6-30 | 1 | No | 4 | No |
| Emotional WB | 4-20 | 11 | No | 4 | No |
| Physical WB | 5-25 | 3 | No | 3 | No |
| Disability-related support | 4-20 | 6 | No | 26 | No |
| FQoL total score | 25-125 | 0 | No | 0 | No |

\*) The total number of participants is 302. This means floor and ceiling effects are present, when 45 people (15%) have the lowest or highest possible score.

### Interpretability

The means and SD of the different subgroups of the baseline data (n = 302) show varied results as illustrated in Table 4.

The mean total score of BC-FQoL scale is higher in male participants. Among age groups, the BC-FQoL is slightly higher in the ≤64 years category. The BC-FQoL scale mean total scores were highest within the ‘able to read and write’ category, followed by the ‘can’t read and write’ category among education groups. From the occupation group ‘merchants’, the total mean score was higher than ‘farmers’ and other occupations. Finally, BC-FQoL total scores are higher among persons affected by leprosy in comparison to persons with podoconiosis. Details of this are presented in Table 5.

**Table 5.**
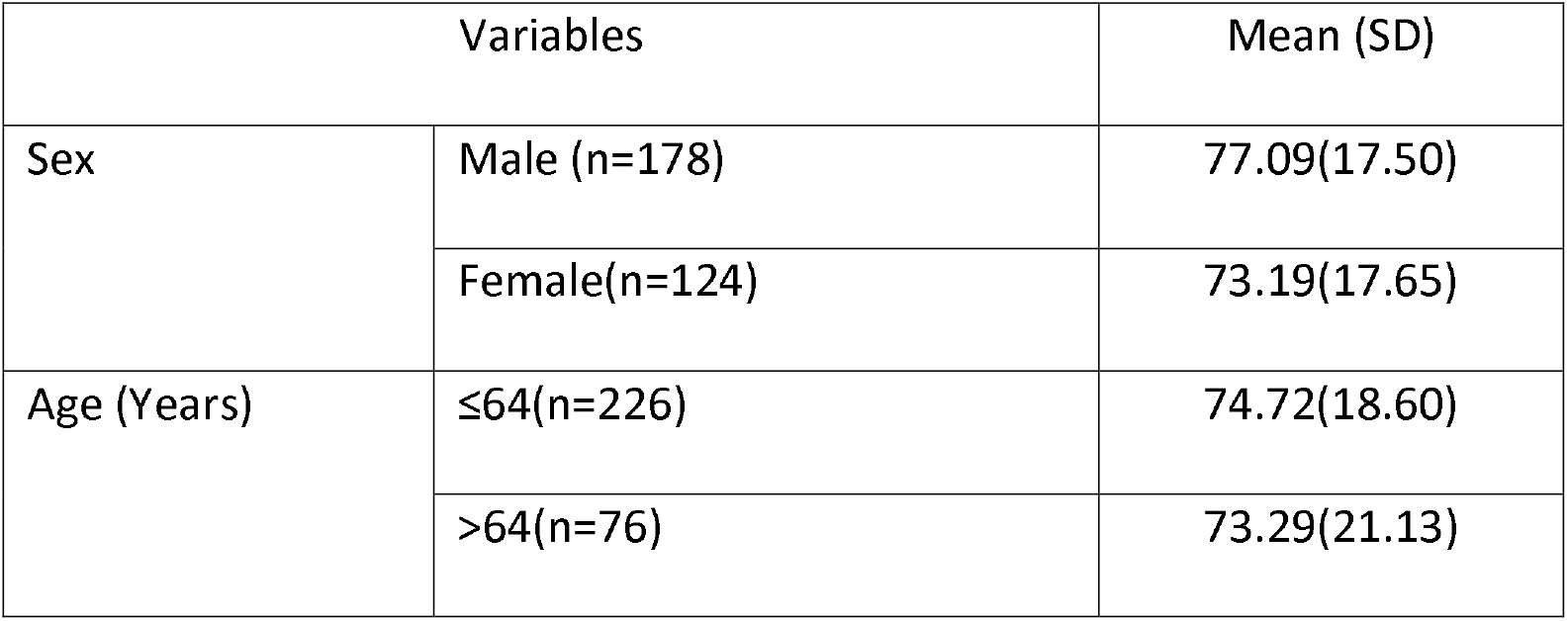
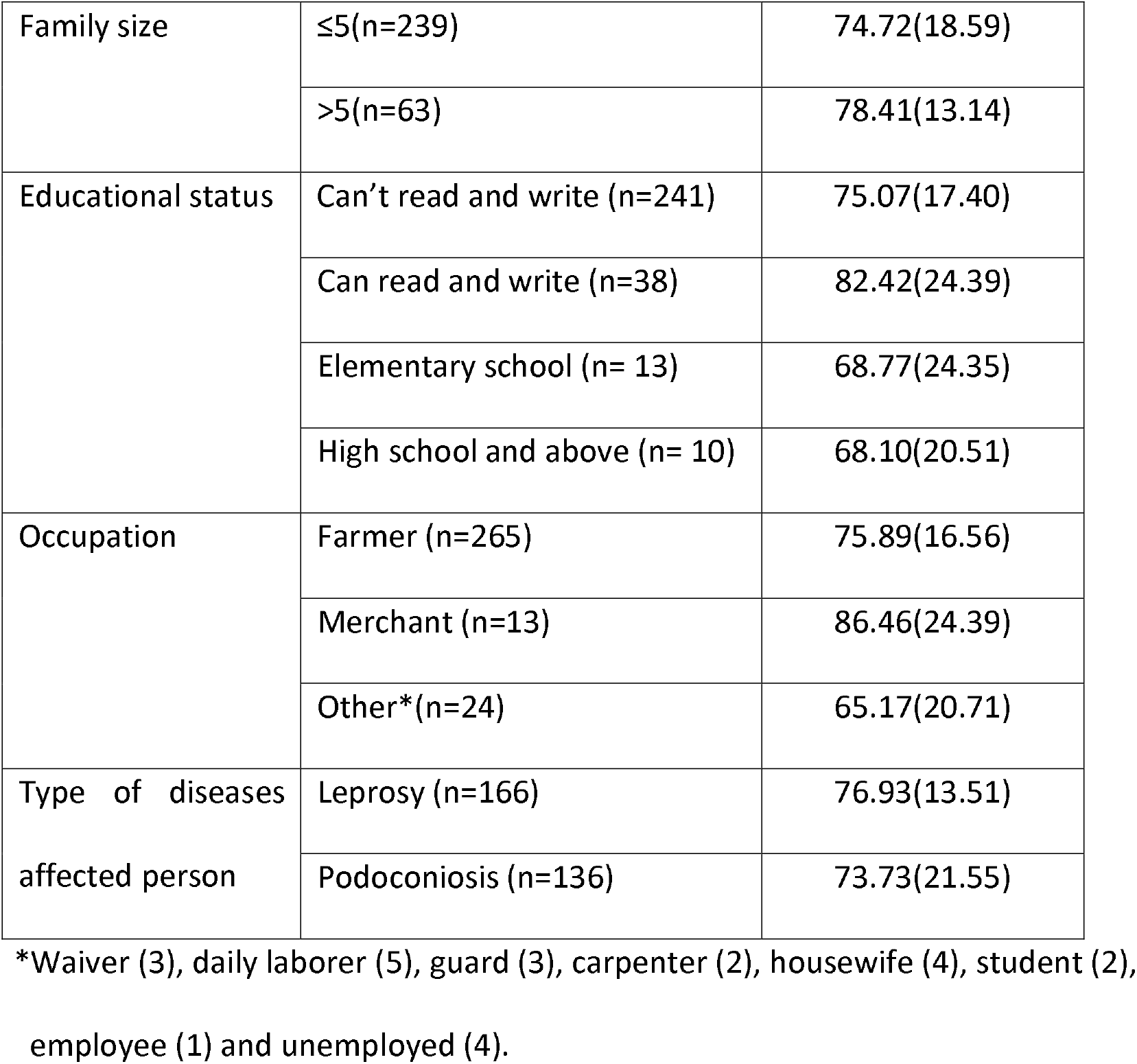
The mean and standard deviation of Beach Centre Family Quality of Life Scale among persons affected by leprosy and podoconiosis.

## Discussion

This study aimed to validate the BC-FQoL Scale cross-culturally into the Amharic language, and subsequently report the cross-cultural validation and psychometric properties of the scale in persons with leprosy and podoconiosis in Northwest Ethiopia.

The cross-cultural adaptation and validation of BC-FQoL Scale was performed based on the recommendations of different scholars [29, 33-35] review. Semantic equivalence was assessed through a process of translation and back-translation. Following these steps, small corrections were made to the Amharic translation of the BC-FQoL scale. This was done based on expert evaluations and the feedback of the original tool developers and input of the target population. The quantitative data analysis results of this study showed a high level of reliability or internal consistency, higher than the cut-off value of 0.7 for Cronbach’s alpha [37, 46]. It was also consistent with the original scale and other researchers’ findings of internal consistency of the BC -FQoL [11, 24, 47-49]. The Cronbach’s alpha of the subgroups was also in the acceptable range in all cases except for the parenting and emotional well-being subgroups, which was nearer to 0.7. Similarly, the result showed an overall excellent agreement or substantial reliability of test-retest ICC of the scale in comparison to different validation studies [37, 38, 50]. This is comparable with a validation study on the Spanish adaptation and validation of the BC -FQoL Scale [49].

The standard error of measurement of the BC-FQoL Scale was <5%, which is considered a very good SEM agreement and is within the acceptable range. This is because of the percentage of the standard error of measurement (SEM) related to the total score of the questionnaire [46, 51]. The SDC of the BC-FQoL Scale was also within an acceptable range, which is in line with findings from other researchers [38, 52-54].

Our CFA indicated that the Amharic version of the BC-FQoL Scale produces reliable results. The first and second order CFA model showed acceptable factor loadings for all items except item 17 (0.47) which has the nearest margin of 0.5. Overall, this result is comparable with the study done by Chiu et al. in China [24], Verdugo et al. in Spain [49] and the original BC-FQoL scale [11]. Similarly, the model fit indices were within acceptable range even though these results came after improving the model by removing item 5, 9 and 25 from the Scale. However, the items which were removed were considered necessary by the experts in the present study, as each item addresses important points in their respective subgroups. For example, item 5 in parenting, which is “my family members help the children with schoolwork and activities”, is vital because of the effect of disability on education [55]. Item 9, “my family members have some time to pursue our own interests”, was also considered essential in the Ethiopian context, which considers giving time for other members of the family an asset of Ethiopian family culture. In addition, item 25, “my family has good relationships with the service providers who provide services and support to our family member with a disability”, has great importance within the Ethiopian context because of the stigma related to disability [55].

Thus the model fit indices results were comparable with the original validation study [11] and other validation studies of the BC-FQoL scale [9, 49]. This study found high composite reliability (CR) which indicated how the subgroup items showed composite reliability to each other within the BC-FQoL scale. This is similar to the study findings with the original scale [11] and Mandarin Chinese versions of the scale results [24].

The convergent validity of this study result indicated an acceptable range even though the average variance extracted (AVE) of the parenting subgroup was 0.494, close to the cut-off value of 0.5. However, this is acceptable on the basis of research done by Fornell and Larcker, which states that if AVE is less than 0.5, and CR is be higher than 0.6, the convergent validity of the construct is still tolerable [56]. This was also the case in our study. Besides the above measure of reliability and validity of the BC-FQoL Scale there were no floor and ceiling effects overall and the subgroups. This has good implications for the scale’s reproducibility and responsiveness [29, 41].

In this validation study, the mean total score of BC-FQoL scale is higher in male participants. This is supported by the study conducted in Ethiopia [57]. Because females engaged more in caregiver stress and not taking on major social roles in education and employment [58]. In addition, Tsutsumi et al. study result showed that an overall lower quality of life score for women than men, a higher mental burden among women compared to men, and perceived stigma affecting QOL of women more negatively than that of men [59]. The BC-FQoL is slightly higher in the ≤ 64 years category. Because above 64 years of age is dependent group of the family in the Ethiopian context [60]. Moreover, the BC-FQoL Scale mean total scores were highest within the ‘able to read and write’ category. This result supported adults with disabilities tend to be poorer than those without disabilities, but education weakens this association [61]. Finally, BC-FQoL total scores are higher among persons affected by leprosy in comparison to persons with podoconiosis. This finding supported by the study done in Ethiopia [57] persons affected by podoconiosis more stigmatized and discriminate than persons affected by leprosy. Persons affected by leprosy were association members that gave chance of getting support from the peers and this provide sense of empowerment [62]. On the other hand, association members of associations of persons affected by leprosy have a chance to take a loan from the association. This supported by research conducted by Wang et al, which indicated that family income is associated with family quality of life [63].

One of the limitations of this study was the use of a convenience sampling technique. Use of the instrument in a more heterogeneous and representative sample of families with disabilities receiving disability management intervention may be needed to further validate the scale. Other studies on the use of the Amharic version of the BC-FQoL are needed in other Amharic-speaking populations and on families of disabled children or families of adults without disabilities, to support its use in other populations. The study used a cross-sectional design to validate the BC-FQoL.

## Conclusion and Recommendation

The Amharic version of the BC -FQoL is reliable and valid in families of persons affected by leprosy and podoconiosis in Northwest Ethiopia. The instrument could be applied in clinical practice, service evaluation and research to assess FQoL in Amharic-speaking populations with leprosy and podoconiosis related disabilities which were or are the recipients of family-based disability management intervention.

## Data Availability

Data available from the author and coauthors. It is available when required.

https://www.dmu.edu.et

## Acknowledgments

First, we would like to express our gratitude to Gozamin Woreda health officials, for facilitating the selection of data collectors and writing a cooperation letter. We would like to acknowledge those professionals participated in the translation and back translation process. We would like to thank Dr. Jean Ann Summers, a team member of the Beach Center Family Quality of Life original developer of the scale as evaluating the back translation. Our acknowledgment goes to all leprosy-affected person association leaders of Bichena, Motta, and Shebel Berenta for their cooperation in recruiting, assisting the data collection process in the specified areas. The research team also acknowledges the data collectors and study participants. The deepest appreciation also goes to Ethiopian National Association of Leprosy affected Person (ENAPAL) staff for facilitating the overall process of data collection especially Tesfaye Tadesse and Abiy Abate. We would like to thank Jane Strugar Kolešnik from Disability Studies in Nederland for her help of editing English language.

## Author Contributions

Conceptualization: Anna T. van ‘t Noordende, Moges Wubie Aycheh, Alice Schippers. Data curation: Moges Wubie Aycheh, Nurilign Abebe Moges, Anna T. van ‘t Noordende. Formal analysis: Moges Wubie Aycheh, Anna T. van ‘t Noordende. Funding acquisition: Anna T. van ‘t Noordende, Alice Schippers, Moges Wubie Aycheh. Investigation: Moges Wubie Aycheh. Methodology: Moges Wubie Aycheh, Anna T. van ‘t Noordende, Nurilign Abebe Moges, Alice Schippers. Project administration: Nurilign Abebe Moges. Resources: Nurilign Abebe Moges. Software: Moges Wubie Aycheh, Anna T. van ‘t Noordende. Supervision: Anna T. van ‘t Noordende, Alice Schippers. Validation: Moges Wubie Aycheh, Nurilign Abebe Moges, Anna T. van ‘t Noordende. Writing – original draft: Moges Wubie Aycheh. Writing – review & editing: Moges Wubie Aycheh, Anna T. van ‘t Noordende, Nurilign Abebe Moges, Alice Schippers.

